# Why is team-based hypertension care failing to take hold in Australia? Real-world evidence from primary care

**DOI:** 10.64898/2026.05.25.26354005

**Authors:** Gautam Satheesh, Kaylee Slater, Ritu Trivedi, Eleanor Clapham, Florence Margaret Lopez, Brendan McCormack, J Jaime Miranda, Shiva Raj Mishra, Gregory M Peterson, Mitchell Sarkies, Aletta E Schutte, Niamh Chapman

**Author notes:** Address for correspondence Dr Niamh Chapman, Faculty of Medicine and Health, School of Health Sciences, The University of Sydney, Camperdown, Sydney, NSW, Australia.

## Abstract

**Objective:** Australia’s shortage of general practitioners (GPs) has intensified interest in team-based care for hypertension, involving pharmacists and nurses. This study explored primary care providers’ experiences, barriers, and facilitators related to implementing team-based care in Australia.

**Design:** Qualitative study using semi-structured interviews with primary care providers.

**Methods:** We conducted 51 interviews with GPs (n=24), nurses (n=12), and pharmacists (n=15), purposively selected from diverse primary care settings. Analysis combined deductive coding, informed by the Theoretical Domains Framework and Consolidated Framework for Implementation Research, with inductive thematic analysis to identify emergent themes.

**Results:** Interviews demonstrated a predominantly GP-centred care model, with nurse and pharmacist involvement largely confined to supporting roles, including blood pressure measurement, prescription refills, patient follow-up and counselling. Their contributions were constrained by barriers at both practice (e.g., limited GP support, fragmented communication across providers) and health system levels (e.g., limited financial incentives and restricted reimbursement pathways). Despite their critical role in care planning, nurses described being “hamstrung” by workload and limited direct funding for hypertension-related services. Pharmacists reported unreimbursed blood pressure checks and restricted funding for medication reviews that constrained the sustainability of their hypertension services. Role ambiguity and the absence of standardised protocols on task sharing further limited collaboration, with nurses and pharmacists describing concerns about overstepping professional boundaries. Attitudes towards team-based care ranged from active disregard (outright rejection) to conditional acceptance and occasional active uptake (strong endorsement).

**Conclusion:** Despite clear willingness among nurses and pharmacists to alleviate GP burden, team-based care is rarely implemented in routine practice. Addressing system-level barriers (funding models that incentivise team-based care and standardised treatment protocols that clarify shared workflows), alongside provider-level barriers (stronger awareness and training that normalises task sharing), is critical to support genuine team-based hypertension care in Australia.

**What is already known on this topic:**

- Only 32% of Australians with hypertension achieve blood pressure control, and care remains heavily GP-centred amid a projected shortage of over 8000 GPs by 2030 and worsening rural access.
- Global evidence strongly supports team-based care involving nurses and pharmacists as clinically effective and cost-effective, yet its uptake in Australian primary care remains limited.

**What this study adds:**

- Drawing on 51 interviews with GPs, nurses, and pharmacists, we identify barriers to team-based care at the practice-level (workload, role ambiguity, professional hierarchies) and system-level (misaligned financial incentives, fragmented communication).
- Despite their willingness to contribute more, nurses and pharmacists remain systematically undervalued, with their contributions constrained by attitudes, incentives, and authority that continue to reinforce GP-centred care.

**How this study might affect research, practice or policy:**

- Australia’s 70% blood pressure control target by 2030 will remain out of reach without urgent structural reforms to primary care—blended funding models, standardised task-sharing protocols, clearer scopes of practice, and interoperable digital systems are prerequisites for genuine, person-centred, team-based hypertension care.

## INTRODUCTION

Hypertension overwhelms health systems worldwide and remains the most common condition managed in Australian primary care.(1, 2) In Australia, one in three adults live with hypertension, yet only 52% are diagnosed and 32% achieve adequate blood pressure (BP) control, far lower than in comparable high-income countries, like Canada (68%).(3, 4) Australia’s National Hypertension Taskforce targets 70% BP control by 2030, but this will require substantial primary care reforms.(3)

By 2030, Australia is projected to face a shortfall of over 8,000 general practitioners (GPs), rising to 13,000 by 2035, given the ageing population.(5) Further, 28% of Australia’s population live in rural settings with limited access to care.(6) Global initiatives, including the World Health Organization’s HEARTS technical package for cardiovascular disease prevention, have leveraged team-based care to expand treatment coverage and achieve BP control at scale in over 40 countries.(7, 8) Team-based care refers to a coordinated, collaborative model of healthcare delivery in which physicians and non-physician providers, including pharmacists, nurses, and allied health and other community health workers, work in partnership, sharing responsibilities and tasks according to their expertise to improve hypertension management.(7, 9, 10) When efficiently implemented, team-based care has been proven to improve BP control, medication adherence, timely follow-up, patient satisfaction and clinic attendance.(11–14)

Australia has started prioritising team-based primary care in health system planning, embedding it within national priorities and policy frameworks.(3, 15, 16) Amidst this growing momentum for reform, there is a need to understand how team-based care is implemented in routine primary care practice and the factors shaping it, through the voices of key health system actors: GPs, pharmacists, and nurses.

## METHODS

### Design

This qualitative study used semi-structured interviews with primary care providers to explore experiences, barriers, and facilitators related to the implementation of team-based hypertension care in Australia. The study was designed as a priori within a broader investigation of clinical workflows in primary care hypertension management. Reporting followed the Consolidated Criteria for Reporting Qualitative Research (COREQ; Supplementary File 3).

### Setting and participants

Primary care providers (GPs, practice nurses, nurse practitioners and pharmacists) involved in hypertension care in Australia were recruited using an information power purposive sampling approach to ensure diversity in roles, practice types (primary care practices, community pharmacies, and nurse-led services), and locations (urban, suburban, rural).(17)

Recruitment occurred through advertisements in primary health network newsletters, social media, partner mailing lists, and snowball sampling. All respondents meeting inclusion criteria (currently practising in primary care and involved in hypertension management) were invited to participate via email, and no additional exclusion criteria were applied. Interviewers had no prior personal or professional relationship with participants. Participants were reimbursed AUD 120 per hour.

### Data collection

Semi-structured interviews were conducted between August 2024 and July 2025 via secure videoconference platforms (Zoom, Microsoft Teams) by trained qualitative researchers [Male: GS, Female: KS, EC, RT, FL] with backgrounds including PhD, PharmD, and Master of Global Health, and experience in primary care and health systems research. The interview schedule was informed by the Theoretical Domains Framework (TDF) to explore individual-level perceptions and behaviours related to diagnosis and management of hypertension, with a particular focus on how practitioners utilise team-based care.(18) The interview schedule was developed in consultation with clinicians and hypertension experts but was not pilot tested. Supplementary File 2 provides the interview schedule, framed as part of the broader study. Participants were informed that the study examined experiences and implementation of team-based hypertension care in Australian primary care and were aware that the research was being conducted by investigators interested in primary care and team-based care. Each interview, involving one participant and maximum two researchers, was audio-recorded and transcribed with participant consent, supplemented by field notes. Transcripts were not returned to participants for comment or correction, and no participant feedback on findings was sought. The research team met quarterly to review data and field notes and refined interview schedule iteratively to explore emerging themes. Interviews continued until sufficient conceptual depth and diversity of perspectives were achieved for each provider group.(17) No repeat interviews were conducted.

### Data Analysis

Transcripts were imported into NVivo 15. A deductive coding framework, informed by the TDF and embedded within the COM-B (Capability, Opportunity, Motivation–Behaviour) model was constructed to examine individual-level determinants (e.g., skills, resources, beliefs) of team-based care (Supplementary Table S1).(19) One independent investigator [GS] thematically coded all transcripts, which were reviewed by a second investigator [KS] to ensure analytical consistency. Inductive thematic analysis then identified attitudes towards team-based care (active uptake, passive uptake, neutral, active regard and passive regard). Emergent themes were identified [GS, NC] and reviewed [KS].

To extend analysis beyond individual-level perceptions, identified themes were interpreted in relation to organisational and system-level determinants, and deductively mapped to the Consolidated Framework for Implementation Research (CFIR) domains of Inner and Outer settings to characterise structural, relational and contextual barriers and opportunities of implementing team-based care.(20)

### Reflexivity and positionality statement

The research team included authors with backgrounds in pharmacy [GS, GMP], medicine [JJM], nursing [BMC, FML], allied health [KS], health systems research [KS, AES, SRM, RT, EC], and implementation science [MS, NC]. The multidisciplinary research team brought prior professional and academic interests in team-based care, which may have influenced the study questions and interpretation of findings. To support reflexivity, the interview schedule was developed in consultation with clinicians and hypertension experts, and discussions during manuscript development helped examine explicit and implicit assumptions, challenge interpretations, and attend to divergent views across provider groups.

### Patient and public involvement

Patients and members of the public were not involved in the design, recruitment, conduct, analysis, or interpretation of this study. The study examined the perspectives of primary care providers (GPs, nurses, and pharmacists), rather than patients.

### Ethics approval

University of Sydney Human Research Ethics Committee (2024/HE000528).

## RESULTS

In total, 24 GPs, 12 pharmacists, and 15 nurses across rural, urban, and suburban primary care settings were interviewed (duration 37–80 minutes) (Table 1). Of the 74 providers who initially expressed interest, 23 did not proceed to interview, due to nonresponse and scheduling conflicts.

**Table 1:** Participant characteristics.

|  |  | GPs | Pharmacist | Nurses |
| --- | --- | --- | --- | --- |
| **Number of participants (N)** |  | 24 | 12 | 15 |
| **Female (%)** |  | 71% (n=17) | 58% (n=7) | 93% (n=14) |
| **Area** |  |  |  |  |
|  | *Rural* | 0% (n=0) | 33% (n=4) | 7% (n=1) |
|  | *Urban* | 71% (n=17) | 17% (n=2) | 80% (n=12) |
|  | *Suburban* | 29% (n=7) | 50% (n=6) | 13% (n=2) |
| **Years of experience** |  |  |  |  |
|  | *<10* | 46% (n=11) | 58% (n=7) | 67% (n=10) |
|  | *10–20* | 37% (n=9) | 42% (n=5) | 6% (n=1) |
|  | *>20* | 17% (n=4) | 0% (n=0) | 27% (n=4) |

Results are organised under six themes reflecting attitudes, provider understanding, task-sharing patterns, and barriers to team-based care.

### 1. Attitudes towards team-based care

Providers expressed a spectrum of attitudes, ranging from rejection to conditional acceptance to active support. While some GPs outright rejected the involvement of nurses and pharmacists, many expressed conditional acceptance, delegating selected tasks (e.g., patient counselling) or supporting involvement under specific conditions (pharmacist-led BP measurement as an interim measure if patients have no other means). At the other end of the spectrum, a subset of providers expressed strong confidence and reliance on nurses and pharmacists. Nurses and pharmacists generally occupied a position of passive uptake; they were supportive of team-based care, but their involvement was dependent on factors, such as GP initiation or endorsement and targeted training (Table 2).

**Table 2:**
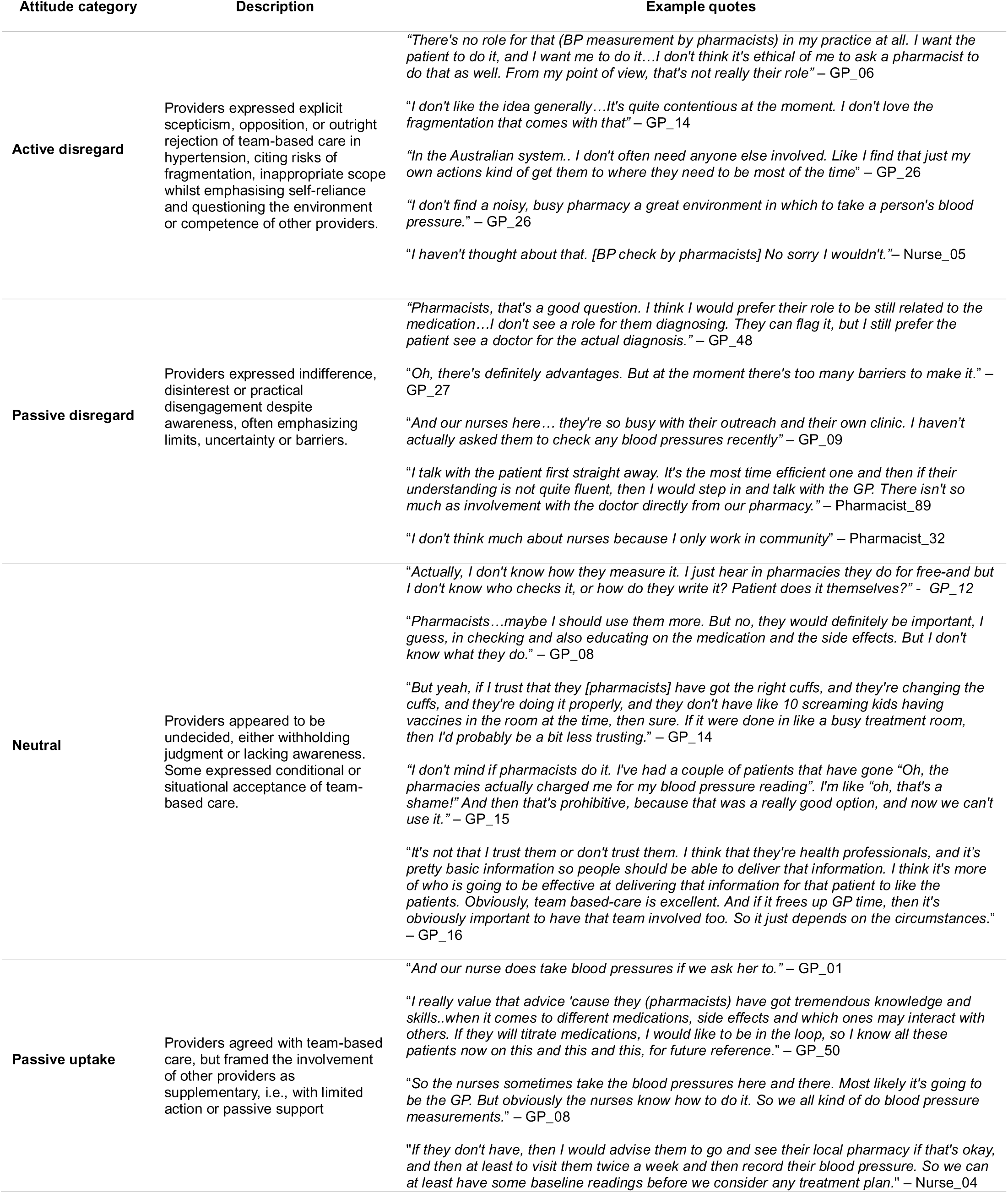
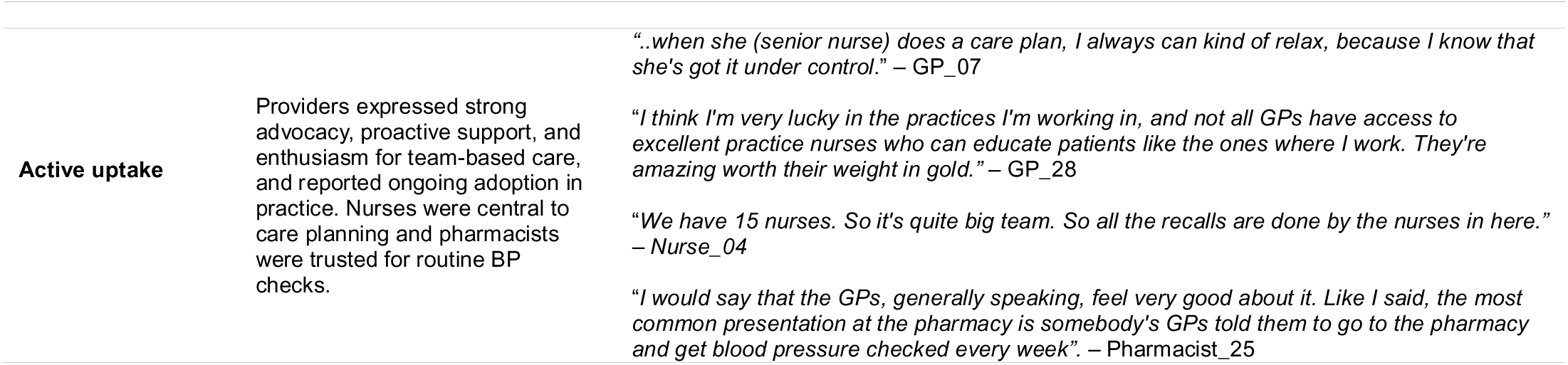
Spectrum of provider attitudes towards team-based hypertension care.

### 2. Current task-sharing and understanding in a largely GP-centred model

There was growing recognition among all provider groups of the importance of team-based care; however, understanding of what it entailed varied widely. Several GPs described team-based care as collaborative care involving other physicians or allied health professionals, without nurses and pharmacists. When GPs described team-based care with nurses, activities centred on BP measurement and patient support. Nurses similarly described GP-centred models, with minimal collaboration between nurses and pharmacists.

As shown in Figure 1, participants described GPs as the primary holders of clinical decision-making responsibilities in hypertension management (diagnosis, treatment initiation and titration), while nurses, and occasionally pharmacists, performed supportive clinical and administrative roles. Nurses reported having to defer to GPs for any changes in management, (e.g., titrating medications), reflecting limited autonomy in routine practice. Similarly, pharmacists noted limited involvement in treatment decisions, with their roles centred on BP measurement and medication counselling without established feedback loops to GPs. Although nurse practitioners have diagnostic and prescribing authority, none reported engaging in either, which remain largely GP-led, suggesting the persistence of established professional hegemonies.

> (on why nurse practitioners do not prescribe) “Patriarchy. Because it’s just this idea of nurse practitioners working in primary care is still a new space and we need to be respectful… I’m just gonna collaborate with them and generally I’m not gonna meet resistance.” – Nurse Practitioner_20

**Figure 1:**
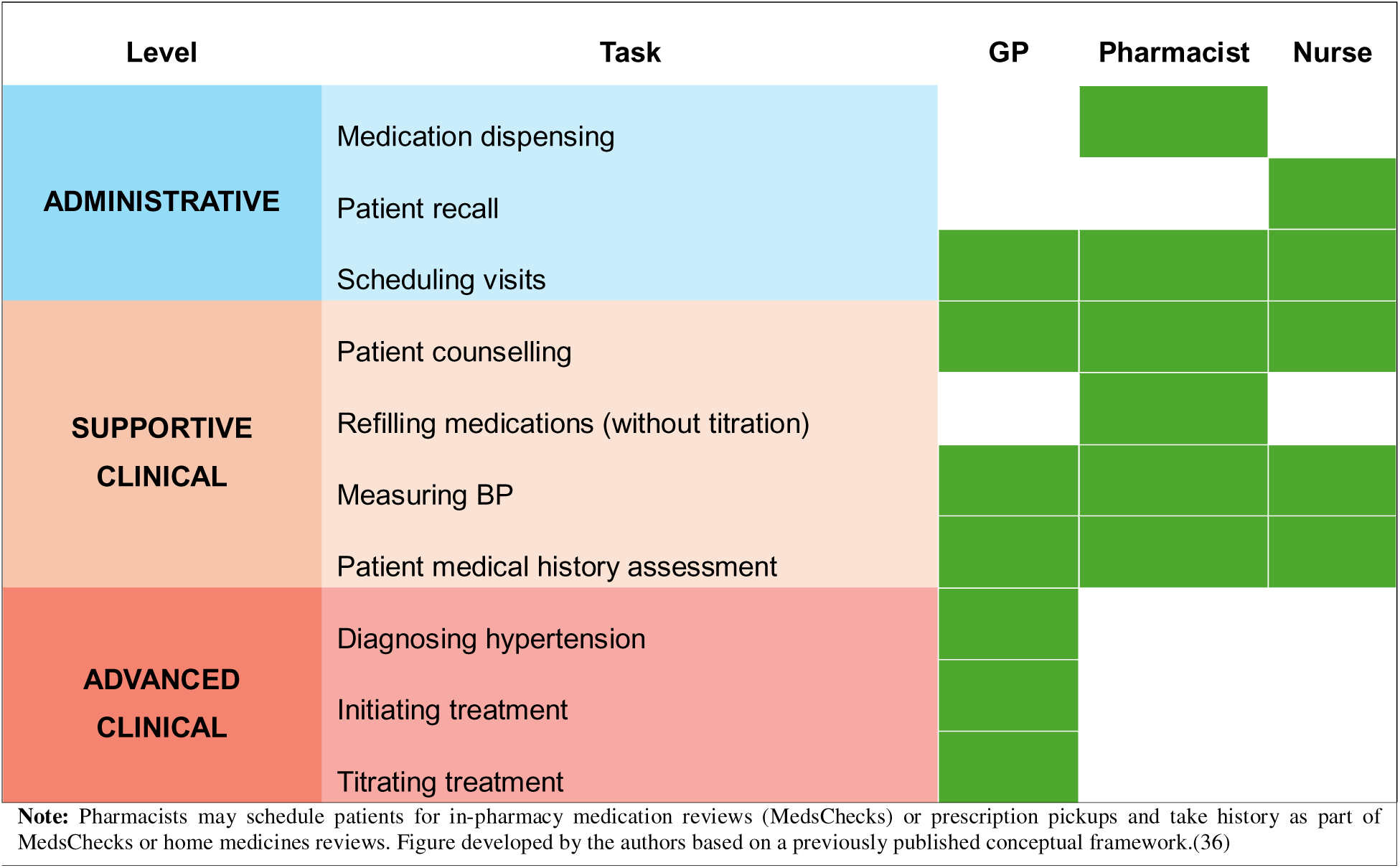
Current scope of task sharing in hypertension care, as reported by provider groups

GPs often emphasised their central role and the trust patients placed in them, even for tasks that could potentially be shared, such as patient counselling.

> “At the end of the day, a GP is very well poised…and has the knowledge and rapport with a patient to be able to provide counselling…I think we’re able to provide a lot more effective counselling and education - we are in a position of trust by the patient.” – GP_16

Some GPs also expressed reluctance in collaborating with specialists and other doctors.

> “…too many cooks spoil the broth” – GP_17

GPs and nurses reported involving allied health providers (e.g., dietitians and exercise physiologists) only when multimorbidity or lifestyle-focused care plans prompted referral.

Participants across provider groups frequently reported existing funding models under Australia’s universal health insurance scheme (Medicare Benefits Schedule; MBS) as favouring GP-centred models and offering limited formal support for task sharing with nurses and pharmacists. A few nurses reported receiving reimbursement for chronic disease services; however, reimbursement is contingent on a GP referral for a care plan, and payments ultimately flowed through to the practice.

### 3. Trusted but structurally hamstrung nurses

This theme highlights the valuable role of nurses in care planning and patient education, while outlining the structural barriers that limit their scope. Participants reported better collaboration in larger clinics with multiple GPs and nurses, wherein structured recall systems facilitated greater nurse involvement in patient follow-up.

However, nurses reported limited capacity to expand their scope due to several systemic barriers, including workload distribution and insufficient financial incentives. GPs reported that nurses were too busy, often heavily involved in aged care, immunisations, and outreach, limiting their role in BP management, which is largely unreimbursed. Nurses similarly reported that practices prioritised activities with existing reimbursements. Further, they stated that rotating rosters and inability to book specific providers undermined continuity of care and limited opportunities to establish familiarity with patients. Nurses themselves articulated frustration at being underutilised despite willingness and experience for more involvement in hypertension management, pointing to structural funding arrangements that reinforce current GP-centred models.

> “I think the system in Australia could be so much better with just a few tweaks… shoving more money to a group of doctors who already cannot take on more patient load… It’s not going to do anything. You should have perhaps engaged a community pharmacist… you should have most definitely utilised your bloody nursing team. They’re already out there in every clinical practice, but totally hamstrung. Utilise the Primary Health Network! The structure’s already there! – Nurse_54

### 4. Accessible yet underutilised pharmacists

GPs and nurses consistently perceived pharmacists as natural collaborators, emphasising that regular patient contact through medication refills offered unique opportunities for team-based care. Pharmacists similarly believed that stronger integration would reduce service fragmentation and improve patient outcomes.

Pharmacists reported that, in practice, their involvement was largely limited to BP measurement and medication reviews for patients with complex needs. BP measurements were conducted mostly at patients’ request and occasionally at the request of GPs, but remained informal and unreimbursed. Most pharmacists offered free BP checks, which were typically reported verbally or recorded in logbooks and handed to patients as no direct communication channels with GPs existed.

Views were not uniform across providers; GPs generally accepted pharmacist-conducted BP readings as a useful interim measure, particularly to rule out anxiety-induced elevated clinic readings (whitecoat hypertension) or when home monitoring was not feasible, although some questioned their validity.

> “Why would I trust that? It doesn’t make any sense to me. What are their readings at the pharmacy? Who they’re talking to? What are their stress levels? What are the other factors?… do it either at home or they do it in the practice. It controls the environment as best as possible, gives us the best quality readings.” – GP_06

Pharmacists expressed that their skills in medication titration were underutilised, citing their recently expanded prescribing role in urinary tract infections. They asserted confidence in prescribing if supported by appropriate training and clear authority. Many reported conducting the government-funded in-pharmacy medication review program “*MedsCheck”*, but noted that rigid eligibility criteria and capped funding limited the sustainability of expanded BP services in busy community settings.

> “…technically, we can only do 20 MedsChecks a month, that’s a cap. But we literally speak to 10 people a day regarding all those issues…” – Pharmacist_23

Unlike nurses, pharmacists reported time constraints only during peak pharmacy hours and described themselves as highly accessible providers who offer walk-ins, longer consultations, and routine opportunistic BP measurement. Pharmacists reported that their potential to support monitoring, education, and adherence was curtailed by perceived GP resistance to expanded roles, especially prescribing.

> “They [GPs] are quite happy to outsource the monitoring. The resistance…the fight is about our ability to prescribe, which, I think in this setting…I can see there would be a lot of backlash from the GPs.” – Pharmacist_38
>
> “Maybe the doctors won’t be happy because they feel that we’re stealing their job away from them. A lot of barriers that must get solved… not easy.” – Pharmacist_39

### 5. Fragmented communication

Nurses and pharmacists consistently reported difficulties in two-way communication with GPs as a key blockage point to collaboration.

> (About communication with GPs) “Efficient two-way? It’s always an efficient one-way! We don’t get any reports back. GP is the central coordinator for all of that. And we are… one pillar of care.” – Pharmacist_38

Community pharmacists, operating as standalone providers and often perceived by nurses and GPs to fragment care, highlighted the absence of integrated digital platforms as a major barrier. Pharmacists mentioned that reliable mechanisms for direct communication or shared workflows remain absent, although national digital health records like “*My Health Record*” hold promise. Most pharmacists noted they almost never received responses following medication review reports and identification of prescription errors. Medication changes were sometimes visible at subsequent prescription refills, but occurred without direct correspondence, reinforcing one-sided communication. Nurses and pharmacists reported no direct communication between each other, and existing pathways rely heavily on one-way exchanges via GPs.

Many GPs acknowledged pharmacists’ potential for increased involvement, and expressed need for better referral pathways, including e-referrals. Fragmentation was reported even within the same practice; where even shared software did not guarantee awareness of other providers’ activities or availability.

### 6. Role ambiguity undermining confidence

Ambiguity around professional roles and task sharing was a recurrent barrier to team-based care across all providers. Nurses described how poor documentation and communication barriers (described above) often created confusion about “who is responsible for what”. Pharmacists, meanwhile, feared that their contributions risked being perceived as encroachment into GPs’ territory.

> “If I made a recommendation like that (medication changes) to a patient, it would be very easily misinterpreted by the GP as questioning their judgment or overstepping professional boundaries…I don’t think that would be good for any relationship with the GP to do that in a community pharmacy.” – Pharmacist_41

Nurses similarly warned that acting to full scope without strong GP backing risked undermining patient trust, rather than fostering collaboration. Role clarity varied depending on team structure. Larger multidisciplinary practices were evidently better at task sharing than smaller GP-led clinics. Even where formal arrangements existed, GPs cited time-intensive multidisciplinary planning and limited remuneration as disincentives for task sharing.

> “We literally can’t build the team care arrangement…you’re spending the same amount of time with them that you would have for both. Because I will do 40 min for those care plan appointments, which is quite a long time with the aim of being able to do comprehensive education. But then it’s a bit of a kick in the guts to get like 50% of what I would normally get for that same time.” – GP_07

Confidence to engage in team-based care varied across providers, shaped by organisational culture and scope. Nurses emphasised that GP endorsement in front of patients bolstered their authority and patient trust, while lack of autonomy or micromanagement undermined motivation. Nurses also added that variable training and underinvestment in general practice nursing further contributed to uneven confidence. Pharmacists expressed confidence in patient education and medication titration, with training and authority, and added that nurses were also well positioned to assume greater responsibility in BP management, given their frequent patient contact. Conversely, nurses often voiced caution over pharmacists’ prescribing scope, highlighting the risk of unsafe practices and overstepping GP boundaries.

Figure 2 summarises the barriers and facilitators towards team-based care identified among provider groups, mapped across inner- and outer-setting influences.

**Figure 2:**
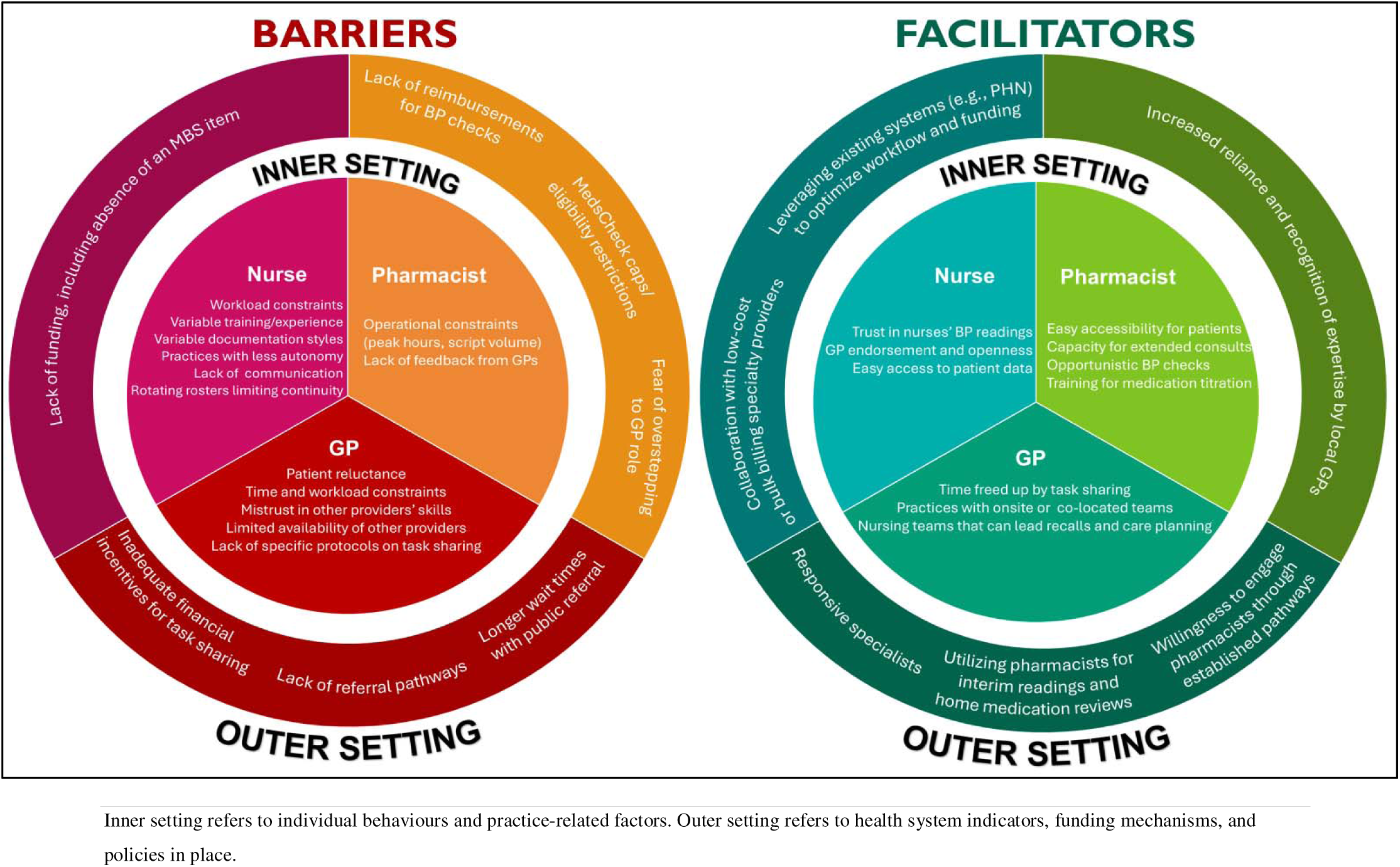
Barriers and facilitators to team-based care across provider groups

## DISCUSSION

This qualitative study highlights that hypertension management in Australia remains predominantly GP-centred, with nurses and pharmacists limited to performing supportive roles in BP measurement, patient follow-up and counselling. Despite general willingness and capability to share tasks and alleviate GP workload, their roles were constrained by individual-, practice- and system-level barriers. In this discussion, we examine the major barriers identified and reforms needed, drawing on lessons from comparable health systems (Table 3).

**Table 3:**
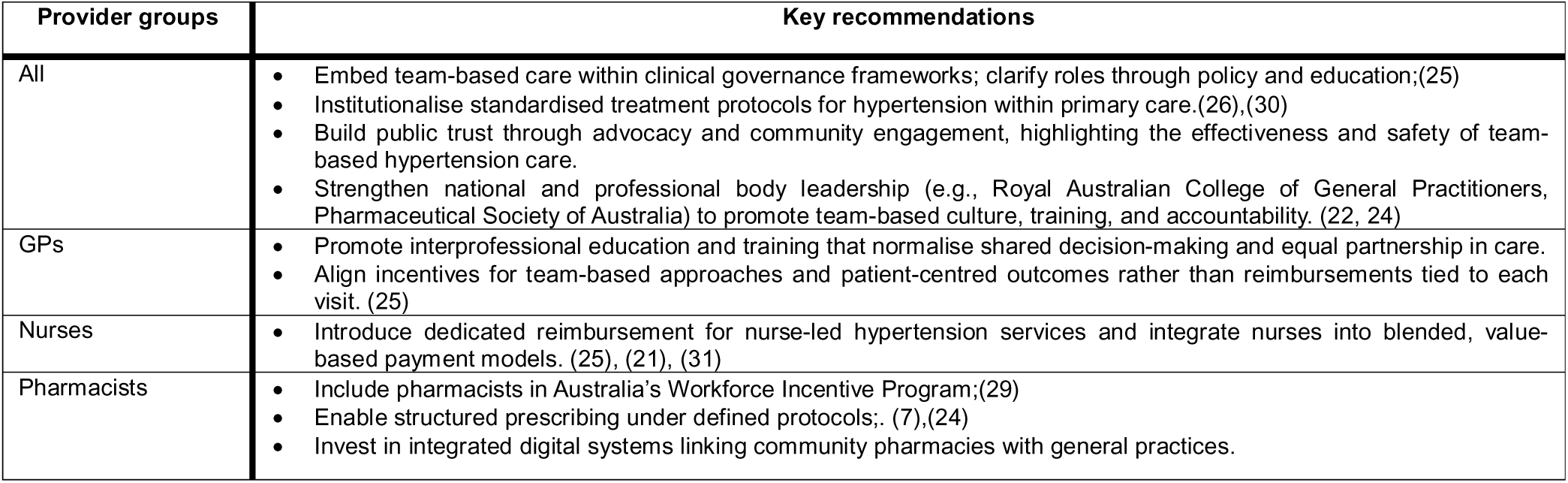
Key recommendations for all provider groups in Australian primary care.

| Provider groups | Key recommendations |
| --- | --- |
| All | • Embed team-based care within clinical governance frameworks; clarify roles through policy and education;(25)<br>• Institutionalise standardised treatment protocols for hypertension within primary care.(26),(30)<br>• Build public trust through advocacy and community engagement, highlighting the effectiveness and safety of team-based hypertension care.<br>• Strengthen national and professional body leadership (e.g., Royal Australian College of General Practitioners, Pharmaceutical Society of Australia) to promote team-based culture, training, and accountability. (22, 24) |
| GPs | • Promote interprofessional education and training that normalise shared decision-making and equal partnership in care.<br>• Align incentives for team-based approaches and patient-centred outcomes rather than reimbursements tied to each visit. (25) |
| Nurses | • Introduce dedicated reimbursement for nurse-led hypertension services and integrate nurses into blended, value-based payment models. (25), (21), (31) |
| Pharmacists | • Include pharmacists in Australia's Workforce Incentive Program;(29)<br>• Enable structured prescribing under defined protocols;. (7),(24)<br>• Invest in integrated digital systems linking community pharmacies with general practices. |

A recent global analysis across 199 countries revealed physician shortage as a structural barrier to hypertension control and reinforced the need for enabling greater involvement of non-physician providers.(14) Our findings suggest that uptake of team-based care in Australia is shaped less by individual capability alone and more by organisational arrangements, funding structures, professional boundaries, and the absence of clear pathways for collaboration across outer (governance, funding, referral pathways) and inner (training, workflow, provider readiness) settings of care delivery. Pharmacists and nurses frequently expressed confidence to contribute more to hypertension care, yet funding models reimburse specific activities rather than patient outcomes, creating misaligned goals. High-quality systems must measure what matters to people, including sustained BP control, rather than task completion.(21)

Narrow and inconsistent conceptualisations of team-based care across providers reflect established professional boundaries, with positional power reinforcing GP-centred models. The ongoing public dispute between national medical associations and pharmacy bodies over pharmacist prescribing exemplifies this resistance to role expansion.(22, 23) The newly implemented national standards to expand nurse prescribing in primary care is likely to provoke similar disputes.(24) Our interviews echoed this tension: even GPs supporting team-based care often described pharmacists and nurses in instrumental terms—“use them” rather than “work with them”— thereby limiting equal partnership.

A recent Australian government ‘Scope of Practice’ review similarly underscored the need to improve role clarity to facilitate collaborative primary care.(25) Hypertension presents a prime opportunity for embedding team-based care in Australia.(14) In other settings, multidisciplinary task sharing in hypertension control programs is generally facilitated by standardised treatment protocols yet to be institutionalised in Australia.(26) Given the strong evidence that team-based care improves BP control, integrating nurses and pharmacists into hypertension management can save thousands of lives annually, representing a highly cost-effective investment in Australia’s health system.(27)

Nurses identified the absence of direct funding for hypertension services as the major barrier, while pharmacists cited unreimbursed BP checks and the monthly cap on funded medication reviews.(28) These concerns have been repeatedly acknowledged in recent government reviews, which have identified Australia’s fee-for-service payment model, where reimbursement is tied to specific activities, as a structural barrier. Transitioning towards blended, ‘value-based’ funding incentivises team-based care by fairly remunerating nurses and pharmacists.(25) Yet, the Australian Government’s Workforce Incentive Program notably excludes pharmacists, contrary to global hypertension control initiatives (e.g., HEARTS technical package) that explicitly advocate for enhanced pharmacist involvement.(29, 30) We note that larger multidisciplinary practices better supported task sharing, compared to smaller GP-run practices. The Strengthening Medicare Taskforce Report (2022) prioritised sufficient funding for general practices to support team-based care.(31)

Australia’s GP shortages and persistently poor BP outcomes warrant substantial funding and workforce reforms.(25) Comparable health systems offer practical lessons: England’s National Health Service introduced the “Additional Roles Reimbursement Scheme” to fund non-physician roles within primary care and help reduce GP workload, providing clear role and task-sharing guidance.(32, 33) In parts of the United States, pharmacists and advanced practice nurses now have prescribing authority. The Canadian Medical Association is advocating for scale-up of publicly insured team-based primary care, currently implemented only in selected provinces.(34) Team-based care is also illustrated by reforms in Latin America enabling nurses and pharmacists to initiate and titrate hypertension treatment, (7) by Australian pilot trials allowing pharmacist prescribing for uncomplicated UTIs,(24) and by Aboriginal Community Controlled Health Organisations.(25, 35) These examples suggest that workforce reform, alongside clear protocols, stronger referral pathways, and more deliberate strategies to strengthen collaboration across provider groups, will be critical in normalising team-based care as Australia’s most sustainable path to improved BP control.

A diverse sample of primary care providers and triangulation across established frameworks strengthened the rigour of our findings.(18–20) A key limitation is the exclusion of other allied health professionals, people living with hypertension as well as priority populations (rural and remote, culturally and linguistically diverse, and Indigenous populations). While we acknowledge their importance in designing equitable care models, they operate within distinct systems and policy environments that warrant separate, dedicated investigation.

## CONCLUSION

Hypertension management in Australia remains predominantly GP-centred, despite clear willingness and capability among nurses and pharmacists to alleviate GP workload and assume greater responsibility. Without substantial workforce and funding reforms that formalise their roles, achieving Australia’s target of 70% BP control by 2030 remains unrealistic. This study identifies provider-level barriers of workload and limited support, alongside system-level financial constraints, offering actionable pathways for sustainable workforce planning, effective task sharing and genuine team-based care. The evidence is unequivocal —hypertension offers a timely and actionable entry point for implementing team-based care in Australia.

## Data Availability

All data produced in the present study are available upon reasonable request to the authors.

## ACKNOWLEDGMENTS

We thank all general practitioners, nurses, and pharmacists who generously shared their time, experiences, and insights to make this study possible.

## DATA SHARING

The de-identified data we analysed are not publicly available, but requests to the corresponding author for the data will be considered on a case-by-case basis.

